# Elucidating the phenotypic variability associated with the polyT tract and TG repeats in CFTR

**DOI:** 10.1101/2020.12.02.20242719

**Authors:** Keith Nykamp, Rebecca Truty, Darlene Riethmaier, Julia Wilkinson, Sara L. Bristow, Sienna Aguilar, Dana Neitzel, Nicole Faulkner, Stephanie Kahan, Swaroop Aradhya

## Abstract

**Purpose:** To evaluate the risk and spectrum of phenotypes associated in individuals with one or two of the *CFTR* T5 haplotype variants (TG11T5, TG12T5 and TG13T5) in the absence of the R117H variant.

**Methods:** Individuals who received testing with *CFTR* NGS results between 2014 and 2019 through Invitae at ordering provider discretion were included. TG-T repeats were detected using a custom-developed haplotype caller. Frequencies of the TG-T5 variants (biallelic or in combination with another CF-causing variant [CFvar]) were calculated. Clinical information reported by the ordering provider (via requisition form) or the individual (during genetic counseling appointments) was examined.

**Results:** Among 548,300 individuals, the minor allele frequency of the T5 allele was 4.2% (TG repeat distribution: TG11=68.1%, TG12=29.5%, TG13=2.4%). When present with a CFvar, each of the TG[11-13]T5 variants were significantly enriched in individuals with a “high suspicion” of CF/CFTR-RD (personal/family history of CF/CFTR-RD) compared to those with very “low suspicion” for CF or CFTR-RD (hereditary cancer testing, *CFTR* not requisitioned). Compared to CFvar/CFvar individuals, TG[11-13]T5/CFvar individuals generally had single organ involvement, milder symptoms, variable expressivity, and reduced penetrance.

**Discussion:** Data from this study provides a better understanding of disease risks associated with inheriting TG[11-13]T5 variants and has important implications for reproductive genetic counseling.

## INTRODUCTION

Cystic fibrosis (CF, OMIM 219700, MedGen 41393), one of the most studied genetic diseases, is caused by biallelic pathogenic variants in *CFTR* and has a highly variable phenotypic spectrum.^1–3^ Pathogenic variants in *CFTR* can alter the abundance, stability, or function of the encoded CFTR protein to varying degrees.^4^ The severity of CF or a CFTR-related disorder (CFTR-RD) is largely dependent on the combination of pathogenic variants and the amount of residual CFTR activity associated with these variants. For example, the most severe pathogenic variants, such as the common Phe508del, eliminate nearly all protein function in the cell. When the residual activity of CFTR is very low (less than 2%) individuals typically have a classic CF phenotype, characterized by multi-organ involvement including severe lung and liver disease, pancreatic exocrine insufficiency, sweat gland dysfunction, and male infertility due to congenital absence of vas deferens (CAVD).^5^ However, when the combination of biallelic pathogenic variants results in residual CFTR activity between 4 and 25%, a range of non-classic phenotypes are often observed, such as CF with pancreatic sufficiency, isolated recurrent pancreatitis, bronchiectasis, chronic sinusitis and/or CAVD in males.^2,6^ In addition to variations in residual activity, genetic modifiers and/or environmental factors can also result in variable expressivity and penetrance of CFTR-related symptoms.^7^

As a class, *CFTR* variants that affect mRNA splicing are often associated with milder symptoms and reduced penetrance due to variable defects in splicing and protein activity. One example of this is illustrated by a set of variants within the polymorphic repeat region of intron 9. This region consists of a thymidine-guanine (TG) repeat element and a thymidine (polyT) tract. The TG repeat in most individuals ranges from 10 to 13 successive TGs, and the polyT tract is most often 9, 7, or 5 uninterrupted thymidines. Importantly, the length of the polyT tract combined with the number of TG repeats is a major determinant of whether exon 10 is included in the CFTR mRNA during splicing.^8–13^ For individuals with a polyT of 9 (T9) and/or 7 (T7) on both chromosomes, greater than 75% of the CFTR mRNA expressed in respiratory epithelial and vas deferens cells includes exon 10 and is full-length (CFTR-fl). The remaining fraction of CFTR mRNA is missing exon 10 (CFTRΔ10) and as a result encodes a non-functional CFTR protein.^14–16^ By contrast, individuals who have a polyT of 5 (T5) on one or both chromosomes have been found to express less than 50% functional CFTR-fl in their cells. Importantly, the proportion of CFTR-fl expressed is further modulated by the TG repeats, such that TG11 adjacent to T5 (TG11T5) results in ∼50% CFTR-fl, TG12 next to T5 (TG12T5) results in ∼25% CFTR-fl, and TG13 adjacent to T5 (TG13T5) is expected to result in <25% CFTR-fl.^14,17,18^

Importantly, decreasing residual activity of the three combinations (haplotypes) of intron 9 repeat variants (TG11T5>TG12T5>TG13T5) correlates with increasing severity and penetrance of CFTR-related symptoms. For example, CF with pancreatic sufficiency (CF-PS) has been reported in ∼38% of individuals with TG13T5 and ∼6% of individuals with TG12T5 (both males and females) with a CF-causing variant (CFvar, < 2% CFTR activity) on the opposite chromosome, but CF-PS has not been reported in individuals who are compound heterozygous for TG11T5/CFvar.^19^ CAVD, along with milder CF-related phenotypes such as idiopathic pancreatitis, bronchiectasis, chronic rhinosinusitis and intermediate sweat chloride levels, has been reported in >95% of males with TG13T5/CFvar, 75-85% of males with TG12T5/CFvar, and 35-55% of males with TG11T5/CFvar.^20,21^ While females with a TG11T5/CFvar genotype are not expected to have CF-PS, ∼5% have been reported to have a milder set of CFTR-related phenotypes as well.^21^ In addition, the common R117H variant (c.350G>A, p.Arg117His) has been found to occur on the same chromosome (in cis) as the TG12T5 variant at a low frequency.^22,23^ While R117H may increase the penetrance and severity of the TG12T5/CFvar genotype, clearly the R117H variant is not required for disease considering the association between TG12T5/CFvar genotype and CFTR-RD has been demonstrated in the absence of R117H.^19^

Considering the relatively high prevalence of the 5T allele (∼10% in the general population), and the wide disparity in clinical outcomes for individuals with TG11T5 compared to those with TG12T5 or TG13T5, it has been recommended that clinical interpretation and reporting of the 5T variant should be done in the context of the number of TG repeats, and irrespective of the R117H variant.^20,24^ In accordance with these recommendations, we custom-developed a bioinformatics haplotype caller to accurately determine the TG-T genotype for each individual who had *CFTR* sequencing performed at Invitae. To date, we have sequenced *CFTR* for more than 500,000 individuals. Here, we report on our findings from these individuals, including assessments of how TG-T genotypes were associated with CF and CFTR-RDs.

## METHODS

### Genetic testing panels and study population

Individuals tested through Invitae between 2014 and 2019 with *CFTR* sequencing results were included in this analysis. *CFTR* sequencing is included on targeted hybrid capture next-generation sequencing (NGS) assays designed for preconception carrier screening and diagnostic testing for certain pediatric disorders and for hereditary cancer across 9 multi-gene panels. In cases where *CFTR* sequencing was included in an assay but not ordered by the referring clinician, the results were not reported, even though the sequencing data was generated.

### Genetic testing and variant interpretation

Genetic testing was conducted using targeted gene panels via next generation sequencing (NGS) as described previously.^25,26^ Briefly, oligonucleotide baits (Agilent Technologies, Santa Clara, CA; Roche, Pleasanton, CA; IDT, Coralville, IA) were used to capture exons plus 10-20 bases flanking intronic sequences and non-coding regions with known clinical implications for genes of interest. Sequencing was performed at an average of 350x coverage (50x minimum). The bioinformatics pipeline integrates community standard and custom algorithms to detect single nucleotide variants, insertions and deletions, structural variants with breakpoints in targeted sequences, and exon-level copy-number variants.

The homopolymer polyT and dinucleotide TG repeat region in *CFTR* sequenced using short read next-generation sequencing (NGS) can be difficult to accurately disambiguate using standard bioinformatics tools. Therefore, an algorithm was developed and validated to specifically call the TG-T haplotypes on both chromosomes for all individuals sequenced for CFTR at Invitae. Interpretation of the TG-T haplotypes and all CFTR variants was performed as previously described.^27^ Briefly, all observed variants were classified as pathogenic (P), likely pathogenic (LP), a variant of uncertain significance (VUS), likely benign (LB), or benign (B) according to the Sherloc variant interpretation framework,^28^ which is based on guidelines from the American College of Medical Genetics and Genomics and the Association of Molecular Pathology (ACMG/AMP).^29^ Pathogenic variants reported in the literature to be associated with CF are referred to as CFvar in the text below. Notably, the T9 and T7 allele have been classified as benign, since they are not associated with disease. The TG10T5 is also classified as benign, while the TG13T5 and TG12T5 variants are classified as pathogenic variants as they are associated with CF-PS or other CFTR-RDs in the majority of individuals with a CFvar on the opposite chromosome. The TG11T5 variant has been classified as pathogenic (low penetrance), a modified classification unique to Invitae, due to a much lower penetrance for CFTR-RDs compared to TG13T5 and TG12T5 alleles.

### CFTR cohorts and data analysis

Individuals for whom *CFTR* sequencing data was available were first divided into those who had requested *CFTR* results in a clinical report (*CFTR* requested) and those who did not (*CFTR* not requested). Review and analysis of de-identified data were approved for waiver of authorization and granted exempt status by the Western Institutional Review Board. The allele frequency (observed number of alleles / total number of sequenced alleles) for each of the TG[11-13]T5 variants was calculated for the total population and each of the two subgroups individually. T-tests were performed to determine if the minor allele frequency (MAF) was different between these two groups.

Two assessments were used to determine whether TG11T5, TG12T5 or TG13T5 were associated with CF or a CFTR-RD. In the first assessment, individuals defined as having a “high suspicion” or “low suspicion” of CF or a CFTR-RD were compared. The “high suspicion” cohort included individuals for whom a personal and/or family history of CF or a CFTR-RD was reported on their requisition form, regardless of whether *CFTR* testing was ordered by the clinician. These individuals were identified in the patient database using a keyword search for the following CFTR-RD symptoms: CF, sweat chloride, immunoreactive trypsinogen, CAVD, meconium ileus. The “low suspicion” group were those individuals referred for hereditary cancer testing and did not request *CFTR* analysis as part of the test panel. All other individuals in the overall study population were excluded from this analysis. Within each of these groups, the number of individuals with each of the following genotype combinations was counted: CFvar/CFvar, CFvar/TG13T5, CFvar/TG12T5, CFvar/TG11T5, homozygous TG13T5/TG13T5, homozygous TG12T5/TG12T5, homozygous TG11T5/TG11T5, TG13T5/TG12T5, TG13T5/TG11T5, TG12T5/TG11T5. While we were unable to determine the phase for the various allele combinations in all individuals in this study through parental testing, it is very likely that the vast majority of individuals in this study with CFvar/CFvar or CFvar/TG[11-13]T5 genotypes have one variant on each chromosome, given that none of the CFvar observed have been reported to be *in cis* with each other or with the TG[11-13]T5 alleles. The common R117H variant (c.350G>A, p.Arg117His) was not considered to be pathogenic (i.e., not a CFvar), and individuals with this variant were excluded from the analysis, due to uncertainty regarding the pathogenicity of this variant alone and the likelihood of a genetic interaction with the TG12T5 variant.^22,23,30,31^ Odds ratios (ORs) comparing the high and low suspicion groups were calculated for each genotype according to the following formula: OR = a*d/b*c, where a is the number of individuals with the genotype in the high suspicion group, b is the number individuals with the genotype in the low suspicion group, c is the number of individuals without the genotype in the high suspicion group, and d is the number of individuals without the genotype in the low suspicion group. Confidence intervals (CIs) and *p*-values were calculated for each OR with the MedCalc’s Odds ratio online calculator (https://www.medcalc.org/calc/odds_ratio.php). Pearson’s chi-squared analysis was used to compare the OR effect across the different genotypes.

Second, in order to better understand the phenotypic variability and severity associated with the CFvar/TG[11-13]T5 genotypes, we identified all individuals in the database who have 2xCFvar or one of the CFvar/TG[11-13]T5 genotypes and counted the number individuals with one or more of the CFTR-related symptom categories listed in Supplemental Table 1. Individuals who “provided indications” included those with symptoms provided at the time of requisition by the referring clinicians or those who self-reported their symptomatic status during a consultation with an Invitae genetic counselor. Individuals were labeled as “No information provided” when the “indication for testing” field was left blank on the test requisition form. All clinical information was provided at the discretion of the ordering clinician on the TRF or of the tested individual at the time of the genetic counseling consultation.

## RESULTS

### Summary of study population

A total of 548,300 individuals had *CFTR* sequencing performed between 2014 and 2019, representing a total of 1.1 million alleles sequenced. *CFTR* genetic testing was requisitioned for 34,195 individuals, and these individuals were younger than those who did not requisition testing for *CFTR*. In the overall population, the majority of individuals were female (78.3%), mean age at time of testing was 47.0 years, and the majority self-reported White/Caucasian ethnicity (61.3%) (Table 1). In this cohort, the minor allele frequency (MAF) of the T5 allele (4.2%) and the distribution of the TG repeats (TG11=68.1%, TG12=29.5%, TG13= 2.4%) was consistent with previous reports in the literature,^20,21^ and similar regardless of whether *CFTR* results were requisitioned for the individual (Table 2).

**Table 1.** Demographic characteristics of study population.

| Characteristic | Total<br>(n=548,300) | CF/CFTR-RD<br>“high” suspicion<br>(n=860) | CF/CFTR-RD “low”<br>suspicion<br>(n=439,331) |
| --- | --- | --- | --- |
| Sex, n (%)<sup>a</sup> |  |  |  |
| Female | 429,414 (78.3) | 447 (52.0) | 369,699 (84.2) |
| Male | 118,871 (21.7) | 413 (48.0) | 69,620 (15.8) |
| Age, years |  |  |  |
| Mean | 47.0 | 24.7 | 52.6 |
| Median | 49 | 25 | 53 |
| Race/ethnicity |  |  |  |
| White/Caucasian | 335,955 (61.3) | 523 (60.8) | 284,579 (64.8) |
| Hispanic | 46,640 (8.5) | 138 (16.0) | 30,776 (7.0) |
| Black/African-American | 34,497 (6.3) | 30 (3.5) | 27,804 (6.3) |
| Asian | 24,019 (4.4) | 37 (4.3) | 16,328 (3.7) |
| Askenazi Jewish | 20,373 (3.7) | 9 (1.0) | 18,183 (4.1) |
| French Canadian | 2,626 (0.4) | 2 (0.2) | 2,412 (0.5) |
| Native American | 1,878 (0.3) | 2 (0.2) | 1,486 (0.3) |
| Mediterranean | 1,636 (0.3) | 3 (0.3) | 1,020 (0.2) |
| Sephardic Jewish | 1,269 (0.2) | 1 (0.1) | 876 (0.2) |
| Pacific Islander | 1,233 (0.2) | 0 | 837 (0.2) |
| Multi-ethnic | 11,632 (2.1) | 23 (2.7) | 9,566 (2.2) |
| Other/Unknown | 66,542 (12.1) | 92 (10.7) | 45,464 (10.3) |

<sup>a</sup>Sex was unknown for 15 individuals in the overall population. Sex was unknown for 12 individuals included in the *CFTR* “low” suspicion group. <sup>b</sup>Race and ethnicity was self-reported by the individuals receiving testing.

**Table 2.**
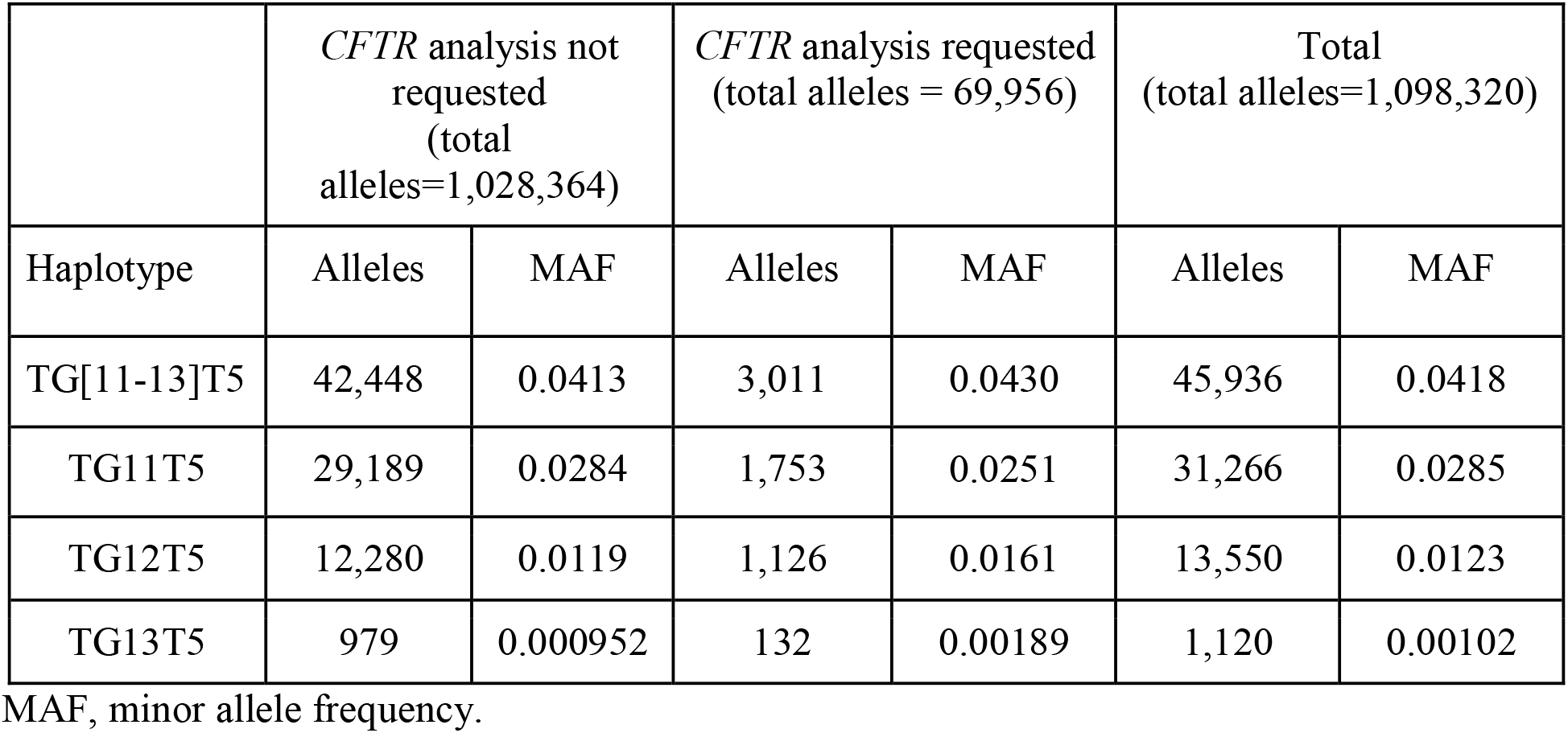
Minor allele frequencies of *CFTR* TG[11-13]5T variants.

### Distribution of T5 alleles in individuals with a high suspicion of a CFTR related disorder

Individuals in the “high suspicion” group were more evenly split among males and females (48.0% vs 52.0%, respectively) compared to the “low suspicion” group (15.8% vs 84.2%). Additionally, age at time of testing was younger in the “high suspicion” group versus “low suspicion” group (mean 24.7 years vs 52.6 years, respectively) (Table 1). When present with a Cystic Fibrosis causative variant (CFvar), each of the TG[11-13]T5 variants were significantly enriched in individuals with a “high suspicion” of CF or CFTR-RD compared to those with very “low suspicion” for CF or CFTR-RD (Table 3). Enrichment was significantly lower (Chi-square statistic, p<0.001) for the CFvar/TG11T5 genotype (OR = 6.9) than for CFvar/TG12T5 (OR = 32.0) and CFvar/TG13T5 (OR = 78.8) genotypes, consistent with reduced penetrance of TG11T5. Interestingly, TG11T5 and TG12T5 homozygotes, along with TG12T5/TG11T5 compound heterozygotes, were absent from the “high suspicion” group, suggesting that these genotypes rarely cause CF or CFTR-RD. By contrast the TG13T5 homozygous genotype was significantly enriched (OR = 255.7) in the “high suspicion” group, similar (Chi-square statistic, p = 0.56) to the 2xCFvar genotype (OR = 581.7).

**Table 3.** Frequency distribution of individuals with CFTR causative variants (CFvar) and TG[11-13]T5 variants for groups with a High Suspicion and Low Suspicion of CF or CFTR-RD

| Genotype | High suspicion<br>(total n=860) | Low suspicion<br>(total n=439,331) | OR (95% CI) | *P*-value |
| --- | --- | --- | --- | --- |
| 2xCFvar | 97 | 96 | 582 (435, 778) | <0.0001 |
| CFvar/TG13T5* | 2 | 13 | 79 (18, 349) | <0.0001 |
| CFvar/TG12T5<sup>§</sup> | 13 | 229 | 29 (17, 52) | <0.0001 |
| CFvar/TG11T5<sup>§†</sup> | 7 | 523 | 7 (4, 15) | <0.0001 |
| Homozygous TG13T5 | 1 | 2 | 256 (23, 2823) | <0.0001 |
| Homozygous TG12T5<sup>§</sup> | 0 | 108 | 2 (0.2, 38) | 0.55 |
| Homozygous TG11T5<sup>§</sup> | 0 | 461 | 0.5 (0.04, 9) | 0.68 |
| TG13T5/TG12T5<sup>§</sup> | 0 | 17 | 15 (0.9, 243) | 0.062 |
| TG13T5/TG11T5<sup>§</sup> | 0 | 11 | 22 (1, 377) | 0.032 |
| TG12T5/TG11T5<sup>§</sup> | 2 | 285 | 4 (0.9, 14) | 0.072 |

\*p<0.05, vs 2xCFvar
<sup>§</sup>p<0.01, vs 2xCFvar
<sup>†</sup>p<0.01 vs CFvar/TG13T5 and CFvar/TG12T5

### Genotype-phenotype associations

Phenotypic variability and severity were assessed in individuals and associated with the corresponding genotype combination when reported by the ordering clinician or self-reported to an Invitae genetic counselor. Of the individuals with a 2xCFvar genotype, phenotype information was provided by the clinician in ∼59% of cases. Phenotype information was provided by the ordering clinician for nearly two-thirds (67%) of individuals with a CFvar/TG13T5 genotype, for 39% of those with a CFvar/TG11T5, and 28% for those with a CFvar/TG12T5 genotype. Overall, 46 individuals within one of the CFvar/TG[11-13]T5 genotypes provided phenotypic information for analysis. In this group, chronic sinopulmonary disease was the most commonly observed category of symptoms, with 14 (30.4%) individuals listing this as a primary indication, which is similar to the percentage of 2xCFvar individuals (28.2%) reporting chronic sinopulmonary complications (Table 4). Recurrent pancreatitis was the next most common phenotypic category for CFvar/TG[11-13]T5 individuals with a total of 10 (21.7%). Interestingly, the majority of these individuals (8/10) had the TG11T5 allele (Table 4), whereas recurrent pancreatitis was very rarely reported (1.3%) for 2xCFvar individuals. Altogether this is consistent with the understanding that mild, CFTR-RD alleles have a higher predisposition for recurrent pancreatitis than classic CF-causative alleles. Eight of the CFvar/TG[11-13]T5 individuals (17.4%) were reported to have intermediate sweat chloride levels (30-60 mEq/L), while none reported elevated sweat chloride levels (>60 mEq/) within the diagnostic range for this lab test. As to be expected, this is in stark contrast to 2xCFvar individuals, in which 28 (18.8%) reported elevated sweat chloride levels and a much smaller fraction (6%) reported intermediate sweat chloride levels. Finally, 2/19 (10.5%) males with a CFvar/TG[11-13]T5 genotype reported obstructive azoospermia or congenital absence of vas deferens (CAVD), which was similar, although slightly lower, than the percentage (14.3%) of 2xCFvar males (6/42) reporting CAVD.

**Table 4.** Distribution of CFTR-related symptoms for TG[13-11]T5 variants.

| **Symptoms** | **2xCFvar** | **CFvar/TG13T5** | **CFvar/TG12T5** | **CFvar/TG11T5** | **Total** |
| --- | --- | --- | --- | --- | --- |
| Chronic sinopulmonary | 42 | 1 | 6 | 7 | 56 |
| Pancreatic insufficiency | 9 | 0 | 0 | 0 | 9 |
| Recurrent pancreatitis | 2 | 1 | 1 | 8 | 12 |
| Gastrointestinal | 22 | 0 | 5 | 0 | 27 |
| Elevated Sweat Cl | 28 | 0 | 0 | 0 | 28 |
| Intermediate Sweat Cl | 9 | 2 | 3 | 3 | 17 |
| Stated CF Dx | 75 | 0 | 3 | 0 | 78 |
| CAVD | 6 | 0 | 2 | 0 | 8 |
| Not affected | 0 | 0 | 3 | 6 | 9 |
| **Patient Responses** |  |  |  |  |  |
| Provided Indications | 149 (59%) | 4 (67%) | 19 (28%) | 23 (39%) | 195 (51%) |
| No information provided | 102 (41%) | 2 (33%) | 50 (72%) | 36 (61%) | 190 (49%) |
| Total individuals | 251 | 59 | 69 | 6 | 385 |

CAVD, congenital absence of the vas deferens; Dx, diagnosis; Cl, Chloride

## DISCUSSION

In this analysis, we describe the significant complexities underlying the clinical interpretation of genotypes involving the *CFTR* TG-T tract based on evidence from the largest study population to date. Because such genomic regions with homopolymers and dinucleotide repeats are often difficult to sequence with NGS and lead to false positive variant calls, we have developed an algorithm that accurately counts both the TG repeat element and polyT tract for each chromosome. This algorithm was validated and applied to the sequencing of more than a half million individuals to ascertain their TG-T genotype. Within this cohort, the MAF of each of the three TG[13-11]T5 haplotype variants was comparable to previous reports.^20,21^ Importantly, we found a significant enrichment of TG[11-13]T5 genotypes in those with a “high suspicion” of CF or CFTR-RD compared to a group with “low suspicion” for CF or CFTR-RD. Enrichment was statistically lower for the CFvar/TG11T5 genotype (OR = 6.9) than for CFvar/TG12T5 (OR = 32.0) and TG13T5 (OR = 78.8) genotypes, consistent with reduced penetrance for TG11T5. Interestingly, TG11T5 and TG12T5 homozygotes were absent from the “high suspicion” group, suggesting these genotypes rarely cause CF or CFTR-RD. By contrast TG13T5 homozygotes were significantly enriched (OR = 255.7) in the “high suspicion” group, and not significantly different from individuals with 2 CF causative variants (CFvar/CFvar, OR = 581.7). Finally, an assessment of reported symptoms in patients with CFvar/TG[11-13]T5 genotypes supports milder symptoms, single-organ involvement, variable expressivity and reduced penetrance for all three TG11T5, TG12T5 and TG13T5 variants. Data from this study provides a better understanding of disease risks associated with inheriting TG[11-13]T5 variants in the absence of the common R117H variant, and has important implications for reproductive genetic counseling. In a recently updated technical standard for CFTR, the ACMG suggested that clinical reporting of the 5T allele in intron 9 should include the number of adjacent TG repeats if possible, given the relatively high frequency of 5T in the population (1 in 10 are carriers) and wide disparity in disease risk associated with TG11T5 carriers compared to TG12T5 or TG13T5 carriers.^24^ In studies reporting on the clinical implications of the TG[13-11]T5 variants,^20^ it is clear that there is significant nuance in their relationship to clinical symptoms and our study provides important clarity for interpreting the clinical significance of the TG-T haplotypes without the presence of the R117H variant. In examining the relationship between TG-T variants and clinical indications for testing, we observed increased risk for disease associated with all three TG[13-11]T5 variants when present with a CF causative variant (CFvar), although to varying degrees. Importantly, individuals with CFvar and TG13T5 or TG12T5 were more likely to report CFTR-related symptoms than individuals with CFvar and TG11T5. Moreover, when TG[13-11]T5 individuals reported symptoms, they were much less severe and more often affecting a single organ (lungs, pancreas, vas deferens) compared to individuals with two classic CF causative variants, consistent with previous reports in the literature. Interestingly, we found that for CFvar/TG11T5 individuals the most commonly reported symptoms are pancreatitis and chronic sinopulmonary disease while CFvar/TG12T5 individuals most commonly report chronic sinopulmonary disease, intermediate sweat chloride levels and a suspected diagnosis of CF. Unfortunately, the total number of individuals with a CFvar/TG13T5 genotype who reported a phenotype was very low (n=4), which limits the spectrum of findings. Nevertheless, these individuals reflect previous reports in the literature with symptoms of chronic pulmonary disease (n= 1), pancreatitis (n = 1), and intermediate sweat chloride levels (n = 2). Altogether, these findings suggest that each allele contributes to a spectrum of CFTR-RDs. Among individuals with clinical information, the observed phenotypic spectrum for those withTG[13-11]T5 alleles illustrates the wide variability and milder severity associated with these alleles compared to individuals with two classic CF causative variants.

While phenotypic analyses of CFvar/TG[13-11]T5 individuals have been reported previously in the literature, there has been very limited information available for individuals who are homozygous or compound heterozygous for the TG[13-11]T5 variants. One case report of a male individual homozygous for TG12T5 demonstrates that this genotype may be associated with CFTR-RD,^32^ specifically recurrent episodes of pancreatitis and elevated sweat chloride levels, but the likelihood that TG12T5 homozygous individuals will present with disease is not clear from this study. With our large population of sequenced individuals, we were able to conclude that TG11T5 and TG12T5 individuals are quite unlikely to report symptoms. Unfortunately, the small number of individuals in the population with a TG13T5 variant limited the number of homozygous individuals available for assessment, although these individuals showed a trend in enrichment similar to patients with 2xCFvar genotypes. Interestingly, although only 2 of 287 TG12T5/TG11T5 individuals reported CFTR-related symptoms at the time of testing (“high suspicion” group), one individual who was not initially included in the “high suspicion” cohort reported chronic sinopulmonary disease upon follow-up with our genetic counseling team. Therefore, clearly the number of patients with CFTR-related symptoms, and corresponding ORs, particularly with milder, mono-organ symptoms, is expected to be higher than observed in this study.

Previous studies in babies identified through newborn screening programs have attempted to measure the penetrance of the TG repeats in causing CF or CFTR-RDs.^19,33^ Other studies have investigated this allele among a population of infertile males.^34^ To our knowledge, this is the first study that investigated a diverse population of individuals with *CFTR* sequencing results available that may or may not be affected by CF or a CFTR-RD. Further, this study includes a broad population of individuals who may have presented with any number of clinical symptoms associated with the full spectrum of CFTR-RDs.

This study is limited by the amount of clinical information provided at the clinician’s discretion on the genetic testing test requisition form or at the individual’s discretion during the genetic counseling consultation to discuss testing results. For example, even when a patient has two CF-causative variants and expected to display symptoms of CF, only 59% of ordering clinicians provided any information about the patient’s disease. The percentage of ordering clinicians who provided phenotypic information for individuals with a CF-causative variant and one of the TG[11-13]T5 alleles was even lower (34%). While this could indicate that many more CFvar/TG[11-13]T5 individuals are asymptomatic than reported, it could also mean that more individuals than expected have CFTR-related symptoms that have been overlooked or simply not provided by the ordering clinician. Importantly, this limitation highlights the challenge with interpreting variants with partial functional activity, and also illustrates the importance of having a patient’s full phenotypic picture when trying to understand variant expressivity and penetrance of mild variants. Finally, although the large number of patients sequenced in this study and their reported phenotypes are useful for better understanding the variable expressivity associated with TG-T alleles, this study population is not necessarily representative of the general population as all included individuals were referred for genetic testing. In order to have more precise understanding of the penetrance and likely course of disease for TG-T carriers, population-based studies will be necessary.

The results from this study complement the recent technical standard from the ACMG^24^ in describing a very large cohort of individuals with *CFTR* sequencing results and describing the spectrum of clinical symptoms observed in individuals with CFvar/CFvar and CFvar/TG[11-13]T5 genotype combinations. The data in this study can help to better counsel individuals undergoing carrier screening or diagnostic testing for the entire spectrum of CFTR-RDs and highlights the need to evaluate these genotype combinations in all individuals tested, regardless of R117H status.

## Data Availability

The data that support the findings of this study are available from the corresponding author upon reasonable request.

