## Supplemental Table 1 for "Elucidating the phenotypic variability associated with the polyT tract and TG repeats in CFTR"

**Supplemental Table 1. CFTR-related categories and associated symptoms**

|  | **CFTR-related symptoms** |
| --- | --- |
| Chronic Sinopulmonary disease | Bronchiectasis |
|  | Respiratory distress |
|  | Recurrent respiratory infections/pseudomonas |
|  | Chronic cough and sputum production |
|  | Chronic wheeze and air trapping |
| Pancreatic disease | Pancreatic insufficiency |
|  | Recurrent pancreatitis |
| Gastrointestinal and nutritional deficiency | Meconium ileus |
|  | Malabsorption |
|  | Rectal prolapse |
|  | Failure to thrive |
|  | Steatorrhea |
| Salt-loss syndrome | Acute salt depletion |
|  | Chronic metabolic alkalosis |
|  | Hyponatremic hypochloremic dehydration |
| Male infertility | Obstructive azoospermia or CAVD |
|  | Non-obstructive azoospermia |
| Labs | Elevated sweat chloride (>60 mEq/L) |
|  | Intermediate sweat chloride (30-60 mEq/L) |
|  | Hypertripsinemia |
| Stated diagnosis (only applied if no symptoms provided) | Suspected Dx of CF |
|  | Clinical Dx of CF |

CAVD, congenital absence of the vas deferens; CF, cystic fibrosis; Dx, diagnosis.
